# Comparative transmission of SARS-CoV-2 Omicron (B.1.1.529) and Delta (B.1.617.2) variants and the impact of vaccination: national cohort study, England

**DOI:** 10.1101/2022.02.15.22271001

**Authors:** Hester Allen, Elise Tessier, Charlie Turner, Charlotte Anderson, Paula Blomquist, David Simons, Alessandra Løchen, Christopher I Jarvis, Natalie Groves, Fernando Capelastegui, Joe Flannagan, Asad Zaidi, Cong Chen, Christopher Rawlinson, Gareth J. Hughes, Dimple Chudasama, Sophie Nash, Simon Thelwall, Jamie Lopez-Bernal, Gavin Dabrera, André Charlett, Meaghan Kall, Theresa Lamagni

## Abstract

**Background:** The SARS-CoV-2 Omicron variant (B.1.1.529) has rapidly replaced the Delta variant (B.1.617.2) to become dominant in England. This epidemiological study assessed differences in transmissibility between the Omicron and Delta using two methods and data sources.

**Methods:** Omicron and Delta cases were identified through genomic sequencing, genotyping and S-gene target failure in England from 5-11 December 2021. Secondary attack rates for Omicron and Delta using named contacts and household clustering were calculated using national surveillance and contact tracing data. We used multivariable logistic regression was used to control for factors associated with transmission.

**Findings:** Analysis of contact tracing data identified elevated secondary attack rates for Omicron vs Delta in household (15.0% *vs* 10.8%) and non-household (8.2% *vs* 3.7%) settings. The proportion of index cases resulting in residential clustering was twice as high for Omicron (16.1%) compared to Delta (7.3%). Transmission was significantly less likely from cases, or in named contacts, in receipt of three compared to two vaccine doses in household settings, but less pronounced for Omicron (aRR 0.78 and 0.88) compared to Delta (aRR 0.62 and 0.68). In non-household settings, a similar reduction was observed for Delta cases and contacts (aRR 0.84 and 0.51) but only for Omicron contacts (aRR 0.76, 95% CI: 0.58-0.93) and not cases in receipt of three vs two doses (aRR 0.95, 0.77-1.16).

**Interpretation:** Our study identified increased risk of onward transmission of Omicron, consistent with its successful global displacement of Delta. We identified a reduced effectiveness of vaccination in lowering risk of transmission, a likely contributor for the rapid propagation of Omicron.

**Funding:** Study funded by the UK Health Security Agency.

## INTRODUCTION

On 11 November 2021, a new SARS-CoV-2 variant, B.1.1.529, was reported in Botswana. Subsequent investigations revealed that the earliest case of this variant was detected on 9 November in South Africa. On 26 November 2021, B.1.1.529 was classified as a ‘variant of concern’ by the World Health Organisation (WHO) and given the designation of ‘Omicron’.^1^ By this point, the variant had most likely already reached several countries.

The Omicron variant has over 30 mutations in the spike protein receptor-binding domain characterised by a minimum of 30 amino acid substructions, three deletions and one mutation.^2-4^ At present, three distinct clades (BA.1, BA.2 and BA.3) have been identified.

The first Omicron case in England, confirmed by genomic sequencing, was detected on 16 November 2021, during a time when the dominant circulating variant was Delta.^5^ A dramatic rise in incidence of COVID-19 ensued, seeing the highest UK incidence reported in the pandemic to date, with over 245,000 cases diagnosed in a single day by late December 2021. Preliminary analyses suggested that the Omicron variant was associated with increased transmission and reduced vaccine effectiveness compared to other SARS-CoV-2 variants.^6^ Data from Gauteng province in South Africa showed a fourth wave of COVID-19 infections with twice as many cases compared to previous waves, indicating rapid spread of this variant.^7^ Surveillance data from England suggested that the Omicron variant may infect more individuals compared to Delta based on the rapid rise in confirmed cases observed following the emergence of the Omicron variant.^8^

Previous emerging SARS-CoV-2 variants, Alpha and Delta, resulted in surges in incidence in England, with increased household clustering observed for both these variants.^9-11^ In contrast, Beta and Gamma variants had lesser impact on case numbers after their emergence.^8^

We applied two different methods (secondary attack rate estimates, household clustering) to assess the transmissibility of the emerging SARS-CoV-2 Omicron variant compared to the Delta variant at a time when both variants were circulating in England.

## METHODS

We assessed transmission of SARS-CoV-2 from cases confirmed as Omicron or Delta variants in England within household and non-household settings, using two different data sources and analytical methods:

1. Secondary attack rates and risk of transmission to named contacts of cases
2. Risk of clusters within cases’ households

### Data sources

#### Cases

All analyses were based on cases in England identified as Delta or Omicron with positive specimen date between 05 and 11 December 2021, a time period where both Omicron and Delta variants were circulating in England.

In accordance with statutory requirements, positive SARS-CoV-2 tests are reported by private and National Health Service (NHS) laboratories to the UK Health Security Agency (UKHSA). Laboratory reports along with self-reported positive rapid Lateral Flow Device (LFD) testing data are stored within the UKHSA Second Generation Surveillance System (SGSS).^12 13 14^

Genomic sequencing in England is co-ordinated by the COG-UK (COVID-19 Genomics UK) consortium and held in the Cloud Infrastructure for Big Data Microbial Bioinformatics database (CLIMB).^15^ SARS-CoV-2 variants were detected from genomic sequencing and genotyping of PCR confirmed cases and by the presence of S-gene target failure (SGTF) identified through PCR processing. Identification of variants is based on UKHSA’s single and multinucleotide polymorphisms variant definitions.^13^ Specimens are selected for sequencing through geographic-weighted population-level sampling of community cases supplemented by targeted selection such as recent international travellers, care homes, or NHS diagnostic laboratories. For these analyses, Delta and Omicron variants were defined through genomic sequencing and genotyping; Omicron cases were also identified by S-gene target failure (SGTF). In England, the CT value threshold for defining SGTF is ≤30CT for both N and ORF1ab genes. Samples may be selected for sequencing or genotyping if they have a ≤30CT value for at least one gene target.

#### Vaccinations

COVID-19 vaccination data for England was obtained from the National Immunisation Management System (NIMS)^16^ system.

#### Contact tracing data

All individuals testing positive for SARS-CoV-2 via PCR were referred for contact tracing with data collated nationally (NHS Test and Trace). Where an individual had multiple positive tests within a 10-day period, only the first positive test triggered the creation of a case record. Information about the case’s symptoms and symptom onset, date of birth, sex, ethnicity, and address were collected during contact tracing. Cases were asked to name people they may have exposed to COVID-19 through close contact during the period they may have been infectious (from two days before symptom onset or, if asymptomatic, test date, until date of contact tracing), with setting type and exposure dates collected. These data were used to assess secondary attack rates in named contacts and to provide proxy information on household size for household clustering analysis.

#### Data linkage and processing

Vaccination status of cases and contacts was obtained by linking case data to the National Immunisation Management System (NIMS)^16^ using a unique patient identifier (NHS number) or combinations of NHS number, forename, first initial, surname, date of birth and postcode. Vaccination status was derived from calculating time between date of vaccination and positive test or symptom onset (cases) or exposure (contacts).

Case data were linked to NHS Test and Trace records using a combination of specimen identifiers, NHS number, and date of birth, to enrich with the number of named household contacts, and to identify individuals with no household contact.

NHS Test and Trace data were enriched with variant information through linkage on specimen identifiers and dates.

#### Transmission to named contacts

For assessment of transmission among named contacts, close contacts named to NHS Test and Trace formed the study population in this analysis. Only contacts of cases with exposure date between 03 and 12 December 2021 were included, capturing the majority of contacts of these exposers and allowing time for follow-up. Close contacts were defined as: household members, face-to-face contacts within one metre of the case, or people within 2 metres for 15 minutes.^17^ Contacts not named by the case (for example, identified as part of contact tracing of international travellers on flights) were excluded. Contacts of cases with missing information on sex were excluded.

The outcome of interest was whether an individual identified as a contact went on to become a secondary case, regardless of their sequencing status. This was identified by matching the named contact data of a COVID-19 PCR-positive case (the exposer) to another COVID-19 PCR-positive case (secondary case) in the NHS Test and Trace data with a symptom onset date (test date if asymptomatic) between 2-14 days (inclusive) after exposure date. Records were matched using forename and surname (including allowing matching of one of these via its initial) and combinations of NHS number, date of birth, postcode, email or telephone number. Where the named contact was a household contact, the symptom onset date (test date if asymptomatic) of their exposer was taken as the exposure date for the contact. To derive the outcome, where multiple contact events (arising either from multiple cases, or the same case on multiple occasions) were matched to a single secondary case record for one individual, rule-based prioritisation prioritising household exposures, and most recent exposures were applied to select a single contact event.

#### Household clustering

The household clustering analyses included ‘index’ cases, the first positive test within a given household. Index cases within a household were laboratory confirmed sequenced cases identified as Omicron or Delta whereas further household cases within any ensuing clusters included cases confirmed by any method (including lateral flow device) regardless of sequencing status to optimise case ascertainment. Index cases included individuals with their first positive specimen during the study period (05 and 11 December 2020) and did not include re-infection cases. Only cases living in private dwellings (including household types: flats, terraced, semi-detached or detached houses or flats) were included.

A household cluster was defined as two or more SARS-CoV-2 cases at the same private residential dwelling, identified through having the same UPRN, within 14 days of the date of first positive specimen. A household forms a cluster on the date the second case tested positive. Sporadic cases refer to a single case detected in a household within 14-day period. To assess household clustering, residential household clusters of COVID-19 were identified from cases’ home addresses self-reported at the time of booking a COVID-19 test, or from the diagnosing laboratory or NHS spine (summary care records). Residential addresses were address-matched against Ordnance Survey reference databases to derive a Unique Property Reference Number (UPRN) to facilitate cluster identification, and Basic Land and Property Unity (BLPU) to identify property type.

Index cases were excluded from this analysis if any of the following applied i) cases from households with a case with an earliest positive test in the preceding 90 days of the index case test date as this could independently reduce the number of susceptible persons in a household ii) household clusters with co-primary cases, defined as more than one case diagnosed within one day of each other, as further household transmission could be from either case iii) cases with no named household contacts at the time of contact tracing by NHS Test and Trace, including those who did not complete contact tracing documents. iv) cases identified through non-community testing i.e. hospital testing (tests captured in Pillar 1), to reduce any bias by including hospitalised patients who would not contribute to household clustering.

#### Descriptive analysis

For transmission among named contacts, contacts of Delta and Omicron cases were described by exposure date, age group, sex, vaccination status and whether they completed contact tracing and by the age group, sex, ethnicity, index of multiple deprivation (IMD) quintile, region of residence, symptom status and vaccination status of their exposer. IMD quintiles for 2019 within England at Lower Super Output Area (LSOA) level (2011 boundaries) were used.

Median serial intervals were calculated for symptomatic index cases and each subsequent symptomatic case among named contacts as the period in days between the dates of symptom onset of the two cases.

The proportions of genotyped and sequenced Omicron and Delta cases with CT value data available that had ≤30CT for both N and ORF1ab genes were assessed to address whether there was any bias in CT values introduced by the use of S-gene status as an indicator for Omicron but not for Delta.

Secondary attack rates, the proportion of close contacts of cases with Omicron and Delta variants that became cases, were calculated. This was based on all cases reported to NHS Test and Trace, including individuals with previous diagnosed infection. For the household clustering analyses, Delta and Omicron index cases were described by specimen test date, age group, sex, ethnicity, IMD, region, number of household contacts and vaccination status. The mean number of household contacts for each variant was described.

### Statistical analysis

To evaluate differences in transmission among named contacts of cases with Omicron and Delta variants, logistic regression models were used to compare the risk of close contacts becoming a case for each variant. Separate models using the same methods were conducted for household and non-household contacts. The multivariable models adjusted for exposure date, characteristics of index cases and contacts (age group, sex, vaccination status), whether the contact completed contact tracing and the symptom status, region of residence, IMD and ethnicity of the index case. To allow for potential differential protection from vaccination, interactions between the variant and the vaccination status of the index case and the contact were also included in the models, and model fit assessed using likelihood ratio tests. To assess any differences in transmission of Omicron compared to Delta in the context of vaccination, adjusted risk ratios of transmission among contacts of cases with Omicron and Delta within exposer and contact vaccination status categories, and adjusted risk ratios of transmission comparing exposer and contact vaccination statuses within contacts of cases with each variant were calculated from these models. Adjusted secondary attack rates and adjusted risk ratios among contacts of cases where both case and contact were unvaccinated were derived from the same models. To support interpretation of the results as risks and changes in risk, adjusted secondary attack rates and risk ratios were calculated, standardised to the study population via post-estimation of subject-specific predicted risks from the models in both Omicron and Delta scenarios. ^18^

For household clustering, to assess whether Omicron cases were more likely to result in clusters of cases at the same residential property compared to Delta, a logistic regression model was conducted. The index case of the household was included in the model with the outcome variable as a binary indicator for clustering.

A multivariable model adjusting for age group, sex, ethnicity, IMD in quintiles, number of household contacts, household type (terraced, semi-detached, detached or flat), earliest specimen date, region, asymptomatic vs. symptomatic and vaccination status prior to infection was carried out. Furthermore, two interaction parameters were assessed to identify any interaction between SARS-CoV-2 variant and the specimen date as well as between variant and vaccination status to account for both changes in PCR test availability in December 2021 and differential vaccine effectiveness by variant, respectively. As for the analyses among named contacts of cases, adjusted risks of clustering and risk ratios of clustering were derived using post-estimation of subject-specific predicted risks from the models in both Omicron and Delta scenarios. ^18 19^ Finally, a further model was constructed based on individuals who had no recent travel history outside of the UK as a sensitivity analysis to consider behavioural and testing differences among who recently travelled.

The household transmission analysis was conducted in Stata version 15.^20^ Transmission among named contacts analysis was conducted in R version 4.0.5. ^21^ The household transmission analysis was conducted in Stata version 15.^20^ Transmission among named contacts analysis was conducted in R version 4.0.5.^21^

## RESULTS

### Secondary attack rates

23,667 Omicron and 59,031 Delta cases with test dates between 05 and 11 December 2021 were linked to contact tracing data; of those, 13,874 Omicron and 40,453 Delta cases had at least one named contact (in any setting) during their contact tracing. After excluding contacts whose exposure dates were outside of the study period, 03 to 12 December 2021, and contacts of cases without recorded sex (n = 54) or IMD (n = 260), 40,123 contacts of Omicron cases and 111,469 contacts of Delta cases were included in the secondary attack rate analysis. They were exposed by 13,680 Omicron and 37,601 Delta cases respectively.

Delta cases reported a mean of 2.0 contacts per case (SD (standard deviation) 2.2), 1.6 (SD 1.5) in the household and 0.4 (SD 1.5) non-household contacts. Omicron cases reported fewer contacts overall, but with more variation; 1.7 contacts per case (SD 2.9) of which 1.1 household contacts (SD 1.3) and 0.6 (SD 2.4) non-household contacts.

Where exposure was in the household, the median serial interval from an exposer to a secondary case was 4 days (IQR (inter-quartile range): 2-6 days) for Delta and 3 days (IQR: 2-5 days) for Omicron.Similar serial intervals were observed for exposure outside the household, with a median 4 days (IQR: 2-7 days) for Delta and 3 days (IQR: 1-5 days) for Omicron.

Of those cases identified by sequencing or genotyping and having CT value data available, 98.7% of Delta cases and 99.2% of Omicron cases had CT≤30 for both N and ORF1ab genes.

The unadjusted secondary attack rate among named household contacts of Omicron cases was 15.0% compared to 10.8% for Delta cases. Similarly, secondary attack rates among named non-household contacts were higher for Omicron than Delta, 8.2% *vs* 3.7%.

Multivariable logistic regression models were fitted with interactions of variant with exposer (index case) vaccination status, and variant with contact vaccination status. These were found to be significant in the household model (p< 0.05), and hence included.

The overall risk ratio of transmission to household contacts of Omicron cases was 1.48 (95% confidence interval [CI]: 1.41 - 1.55) compared to those of Delta cases; for non-household contacts of Omicron cases, this was even greater at 2.14 (95% CI: 1.91 - 2.40) (Table 1). Although the effect of variant on risk of transmission varied according to the vaccination status of both the exposer and contact (Table 1), adjusted secondary attack rates were consistently higher for Omicron than Delta for every stratum of vaccination dose for index cases or contacts.

**Table 1:** Adjusted* secondary attack rates and adjusted risk ratios of transmission to named contacts from Omicron compared to Delta cases in household (1A) and non-household (1B) settings. Exposer and contacts with unknown vaccination status omitted. Household settings

|  | Adjusted secondary attack rate Delta (95% CI) | Adjusted secondary attack rate Omicron (95% CI) | Adjusted risk ratio of Omicron vs Delta (95% CI) | p |
| --- | --- | --- | --- | --- |
| All | 10.5% (10.3% - 10.7%) | 15.6% (15.0% - 16.1%) | 1.48 (1.41 - 1.55) | <0.0001 |
| Contact vaccination status |  |  |  |  |
| Unvaccinated | 12.9% (12.4% - 13.5%) | 15.9% (14.8% - 17.0%) | 1.23 (1.14 - 1.32) | <0.0001 |
| 1 dose + 21 days | 10.6% (9.7% - 11.5%) | 14.8% (12.8% - 16.8%) | 1.40 (1.19 - 1.63) | <0.0001 |
| 2 doses + 14 days | 11.2% (10.8% - 11.6%) | 18.3% (17.4% - 19.3%) | 1.64 (1.55 - 1.75) | <0.0001 |
| 3 doses + 14 days | 7.6% (6.9% - 8.3%) | 16.1% (14.5% - 17.7%) | 2.13 (1.87 - 2.43) | <0.0001 |
| Exposer vaccination status |  |  |  |  |
| Unvaccinated | 11.8% (11.4% - 12.3%) | 15.8% (14.7% - 17.0%) | 1.33 (1.23 - 1.44) | <0.0001 |
| 1 dose + 21 days | 9.6% (8.8% - 10.4%) | 14.9% (13.1% - 16.8%) | 1.55 (1.34 - 1.81) | <0.0001 |

1A: Household
|  | Adjusted secondary attack rate Delta (95% CI) | Adjusted secondary attack rate Omicron (95% CI) | Adjusted risk ratio of Omicron vs Delta (95% CI) | p |
| --- | --- | --- | --- | --- |
| 2 doses + 14 days | 10.0% (9.6% - 10.3%) | 15.8% (15.1% - 16.5%) | 1.58 (1.50 - 1.67) | <0.0001 |
| 3 doses + 14 days | 6.2% (5.3% - 7.1%) | 12.4% (10.9% - 13.8%) | 1.99 (1.66 - 2.39) | <0.0001 |

\* with additional adjustment for exposure date, characteristics of index cases (age, sex, IMD and ethnicity, symptom status, region of residence) and contacts (age group, sex, whether they completed contact tracing).

1B: Non-household
|  | Adjusted secondary attack rate Delta (95% CI) | Adjusted secondary attack rate Omicron (95% CI) | Adjusted risk ratio of Omicron vs Delta (95% CI) | P |
| --- | --- | --- | --- | --- |
| All | 3.9% (3.7% - 4.2%) | 7.6% (7.2% - 8.1%) | 2.14 (1.91 - 2.40) | <0.0001 |
| Contact vaccination status |  |  |  |  |
| Unvaccinated | 5.1% (3.4% - 6.8%) | 10.1% (7.4% - 12.8%) | 1.99 (1.32 - 2.99) | 0.0009 |
| 1 dose + 21 days | 6.0% (3.2% - 8.9%) | 7.4% (3.8% - 11.0%) | 1.22 (0.62 - 2.39) | 0.560 |
| 2 doses + 14 days | 5.9% (5.1% - 6.7%) | 9.6% (8.4% - 10.9%) | 1.64 (1.43 - 1.87) | <0.0001 |
| 3 doses + 14 days | 3.0% (2.2% - 3.8%) | 7.3% (5.7% - 9.0%) | 2.43 (1.80 - 3.29) | <0.0001 |
| Exposer vaccination status |  |  |  |  |
| Unvaccinated | 4.9% (4.0% - 5.8%) | 8.8% (7.2% - 10.4%) | 1.33 (1.23 - 1.44) | <0.0001 |
| 1 dose + 21 days | 4.7% (3.2% - 6.3%) | 6.7% (4.6% - 8.9%) | 1.55 (1.34 - 1.81) | <0.0001 |
| 2 doses + 14 days | 3.7% (3.4% - 4.1%) | 7.5% (6.9% - 8.0%) | 1.58 (1.50 - 1.67) | <0.0001 |
| 3 doses + 14 days | 3.1% (2.1% - 4.2%) | 7.1% (5.7% - 8.4%) | 1.99 (1.66 - 2.39) | <0.0001 |

\* with additional adjustment for exposure date, characteristics of index cases (age, sex, IMD and ethnicity, symptom status, region of residence) and contacts (age group, sex, whether the completed contact tracing).

For Delta cases, adjusted secondary attack rates were lowest among exposers and contacts with three doses of vaccine in both settings (Tables 1a and 1b). For contacts exposed to Delta, secondary attack rates were 3.0% (95% CI: 2.2% - 3.8%) for individuals who had received 3 doses *vs* 5.1% (95% CI: 3.4% - 6.8%) in unvaccinated contacts in non-household settings, and 7.6% *vs* 12.9% in household settings. Similarly, transmission was reduced according to number of vaccine doses received by the exposer, 3.1% (95% CI: 2.1% - 4.2%) *vs* 4.9% (95% CI: 4.0% - 5.8%) for three doses vs no doses in non-household settings and 6.2% (95% CI: 5.3% - 7.1%) *vs* 11.8% (95% CI: 11.4% - 12.3%) in household settings. The adjusted risk ratio of transmission for contacts who had received a third vaccine dose compared to two doses was 0.51 (95% CI: 0.39 - 0.66) for non-household cases and 0.68 95% CI: (0.562 - 0.74, p <0.0001) for household contacts (Supplementary Table 1B).

The impact of exposer or contact vaccination on transmission rates for Omicron cases was considerably attenuated compared to Delta, with secondary transmission rates similar for household contacts regardless of number of vaccine doses received, although marginally lower for exposers with 3 doses (12.4%) compared to less than 3 doses or unvaccinated groups (14.9 to 15.8%; Table 1). In non-household settings, a gradient of decreasing secondary attack rates was evident for Omicron cases according to increasing number of vaccination doses received by exposers, although less distinct for number of vaccination doses received by contacts. Adjusted risk ratio analyses showed however a clear protective effect in contacts (0.88, 95% CI: 0.79 - 0.97, p=0.0129) or exposers (0.78 (95% CI: 0.69 - 0.88), p <0.0001) having received 3 doses (compared to 2 doses) in household settings (Supplementary Table 1B). In non-household settings, a protective effect for contacts having received 3 doses vs 2 doses was observed (0.76 (95% CI: 0.61 - 0.94), p=0.0115), but there was no evidence of differences in protection according to number of doses received by exposers (Supplementary Table 2).

Adjusted secondary attack rates in households were slightly higher for unvaccinated contacts of unvaccinated Omicron cases (16.2%, 95% CI: 14.8% - 17.6%) compared to unvaccinated Delta cases (14.6%, 95% CI:13.9% - 15.3%) with an adjusted risk ratio of 1.11 (95% CI: 1.01 - 1.22). Among unvaccinated non-household contacts of unvaccinated cases the difference between secondary attack rates for Omicron and Delta was more marked, with 11.6% (95% CI: 8.2% - 14.9%) for Omicron and 6.3% (95% CI: 4.1% - 8.6%) for Delta and an adjusted risk ratio of 1.84 (95% CI: 1.19 - 2.85).

Compared to contacts aged 30-39 years, risk of household transmission was generally lower in all other age groups (except 40-49 years old), particularly children (Figure 1). Likelihood of transmission was also lower in male than female contacts. Where assessed according to characteristics of the exposer, exposers aged under 30 were less likely to transmit infection to their contacts than exposers aged 30-79 years old. Similarly, in settings outside the household, exposers aged under 20 were less likely to transmit than 30-69-year-olds. Non-household contacts of exposers in London were more likely to become cases than those in the reference region (East Midlands) and household contacts of exposers in the North West were less likely to become cases. Within households, cases who reported being asymptomatic (13.5% of Delta, 8.8% of Omicron) were half as likely to transmit to their household contacts (aOR 0.47 (0.44-0.51)) than those reporting symptoms, but no such (significant) differences were seen for non-household exposures.

**Figure 1:**
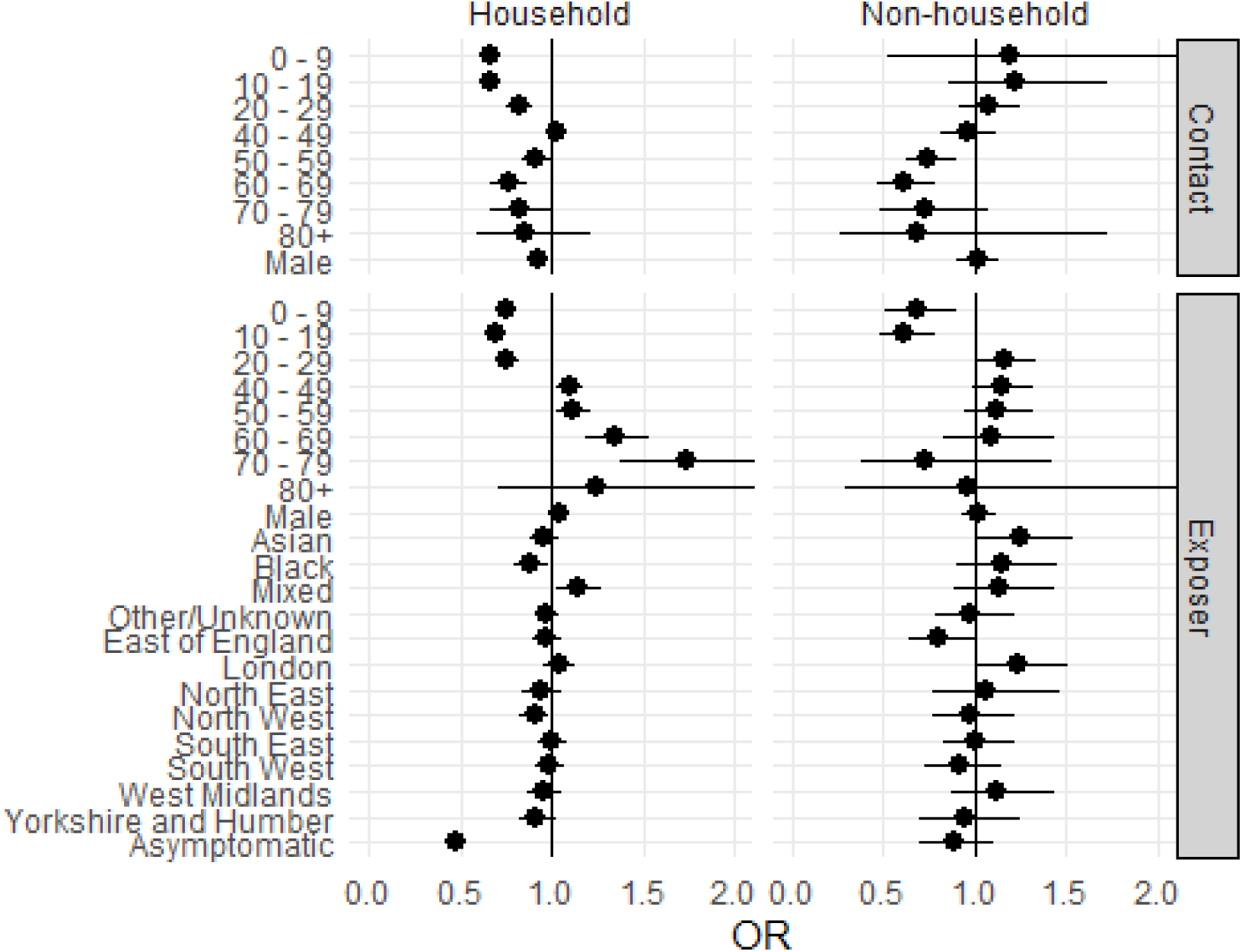
Transmission to named contacts: adjusted odds ratios for selected variables* from multivariable analyses (x-axis limited to 2), 05 to 11 December 2021, England *with additional adjustment for variant, exposer vaccination status, contact vaccination status, interaction of variant with exposer vaccination status, interaction of variant with contact vaccination status, whether contact completed contact tracing, exposer IMD quintile, date of exposure. Missing values omitted for all categories.

**Figure 2.**
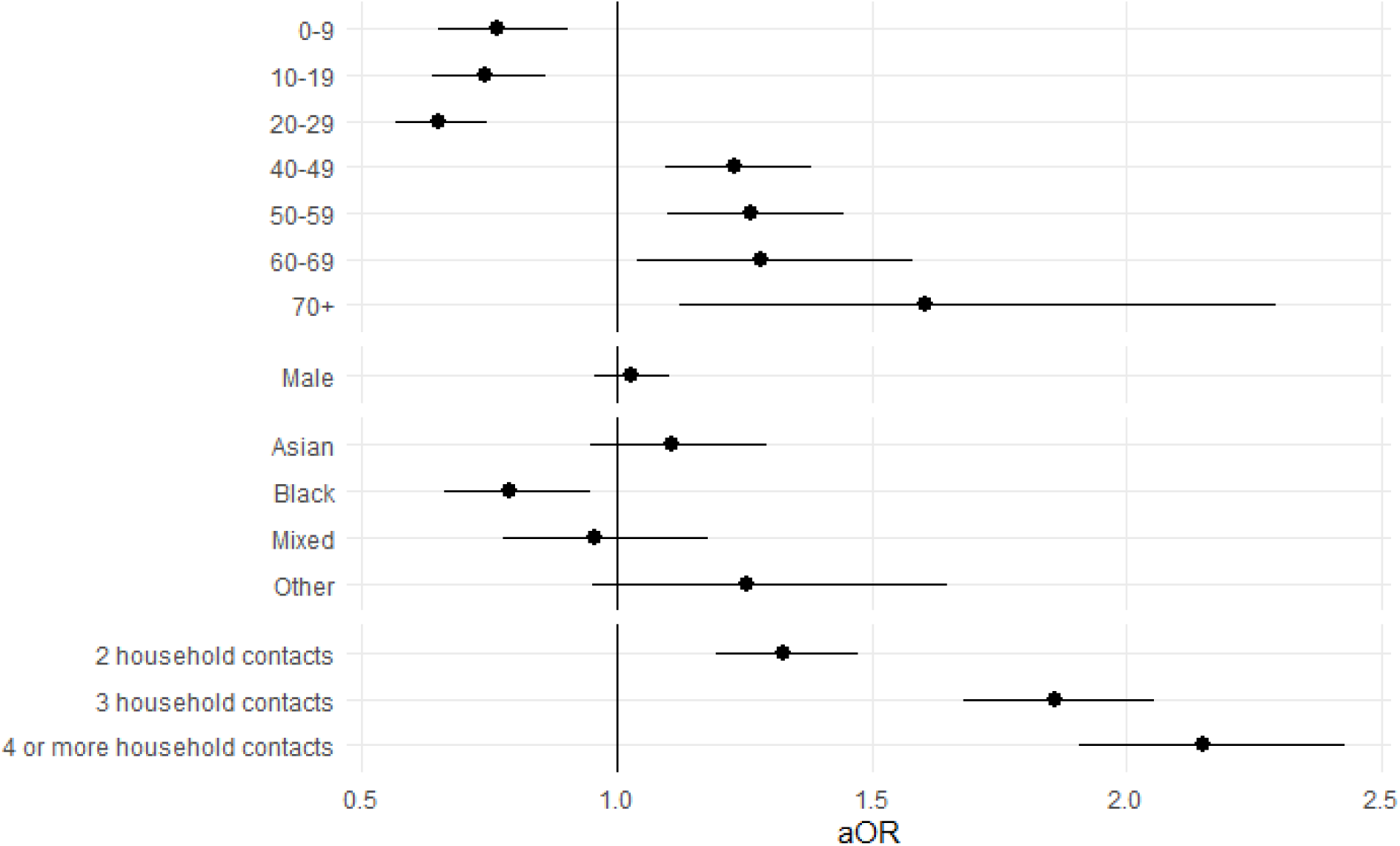
Household clustering for selected variables from multivariable analyses: adjusted odds ratios, 5 to 11 December 2021, England* *The full adjusted model includes adjustment for variant (Omicron and Delta), sex, age group, ethnicity, IMD, household type, earliest specimen date, region, vaccination status, number of household contacts, symptomatic status.

### Household clustering

For inclusion in the household clustering analysis, a total of 307,034 individuals tested positive for SARS-CoV-2 for the first time of which 60,393 cases were confirmed as Delta by genotyping or sequencing and 21,402 identified as Omicron through genomic sequencing or the presence of S-gene target failure, representing a total of 26.6% of all cases reported England during the study period, 05 to 11 December 2021.

After exclusion criteria were applied, 8,692 Omicron and 29,094 Delta index case were included in the household clustering analysis. The mean number of named household contacts was 1.96 (range 1-11) and 2.37 (range 1-10), respectively for Omicron and Delta cases.

Of the cases included in the analysis, 16.13% (1,404) Omicron cases resulted in household clustering, compared to 7.34% (2,136) Delta cases.

The multivariable logistic regression model showed a significant effect modification between variant and the vaccination status of the index case and this interaction was retained in the final model.

Postestimation analysis to assess the risk ratio of household clustering found that the overall risk ratio was 3.54 (95% CI (3.29 to 3.81) for Omicron compared to Delta variants. Furthermore, for each of vaccination status there was an increased risk of household clustering for Omicron compared to Delta variants, most notably among index cases who were ≥14 days post their third vaccine dose with an risk ratio of household clustering of 6.81 (95% CI (4.91 to 9.46) (Table 2).

**Table 2.** Risk of household clustering for Omicron vs. Delta by vaccination status of the index case.

| Vaccination status of index case | Adjusted risk of clustering, Delta | Adjusted risk of clustering, Omicron | Adjusted risk ratio (aRR)<br>Omicron vs Delta |
| --- | --- | --- | --- |
| Overall | 6.58 (6.31 to 6.86) | 23.32 (21.97 to 24.67) | 3.54 (3.29 to 3.81) |
| Unvaccinated | 7.79 (7.14 to 8.45) | 24.83 (22.04 to 27.62) | 3.19 (2.79 to 3.64) |
| $\geq 21$ days post dose 1 | 6.10 (4.98 to 7.22) | 24.93 (20.55 to 29.31) | 4.09 (3.19 to 5.23) |
| $\geq 14$ days post dose 2 | 6.19 (5.73 to 6.64) | 22.02 (20.55 to 23.5) | 3.56 (3.25 to 3.90) |
| $\geq 14$ days post dose 3 | 3.05 (2.13 to 3.98) | 20.80 (17.64 to 23.97) | 6.81 (4.91 to 9.46) |
| Unknown vaccination status | 6.80 (5.35 to 8.25) | 27.54 (22.49 to 32.59) | 4.05 (3.06 to 5.37) |

The effect modification between specimen date and variant was evaluated and identified as not being significant (p=0.123).

Additional factors associated with likelihood of household clustering were age, with younger (<30 years old) index cases having the lowest likelihood of resulting in household clustering, compared to 30-39 years old, and those over 40 years old having higher risk, and ethnicity, with reduced clustering for black vs white index cases.

The overall results of the model for household clustering among Omicron vs Delta index cases did not significantly change in a restricted model excluding those with travel history outside of the UK, showing an overall adjusted odds ratio of 4.53 (95% CI 4.04 to 5.08)), compared to 4.51 (95% CI 4.03 to 5.05) in the final model including those with travel history.

## DISCUSSION

Our study confirms early observations suggesting increased transmission for Omicron compared to Delta.^8^ We identified a significant attenuation of the protective effect of a booster dose of vaccine in reducing risk on onwards transmission from Omicron cases, or susceptibility to infection in contacts, a likely contributor to its rapid expansion in the UK and other countries worldwide. The UK experienced a vertiginous acceleration in incidence since the arrival of Omicron on its shores in November 2021, breaching 2,000 reported cases per 100,000 population by 04 January 2022. The swift domination of Omicron globally highlights the importance of rapid genomic surveillance to detect and better understand the impact of new variants on disease incidence, hospitalisations and mortality.

The emergence of Omicron triggered several public health measures to be reinstated (‘Plan B’). These included compulsory face coverings in most public indoor venues other than hospitality, the implementation of NHS COVID Passes showing proof of vaccination for specific settings, and return to work from home, where possible.^22^ Coupled with evidence of reducing protection from waning immunity, public campaigns strongly encouraged a third dose (booster) of vaccine, which saw the public queuing for hours to receive vaccination ahead of our fourth wave. Our assessment of the impact of this third dose for either cases or their contacts indicates modest benefit in reducing transmission, although a clear and substantial benefit to patient outcomes and maintaining a functioning health service was likely to have been achieved through the booster immunisation programme.^23^

Our study utilised complementary but distinct methods to assess transmission to named contacts identified through contact tracing, and household clustering through geocoding of surveillance data. Our results suggest that the Omicron variant is far more transmissible than the Delta variant in both households and non-household settings. Our findings support those of Gu *et al*. who reported probable Omicron transmission between two fully vaccinated travellers staying in rooms across the corridor, despite quarantine restrictions.^24^ The elevation in transmission risk between Omicron and Delta was particularly apparent in non-household settings, with twice the likelihood of contacts developing infection than for Delta; this increased risk was particularly notable in transmission to unvaccinated non-household contacts of unvaccinated Omicron cases (compared to their Delta counterparts). This suggests that briefer, less proximate contact events may be sufficient for transmission from Omicron cases (relative to Delta). It could also, however, relate to milder symptoms for Omicron than Delta, leading to reduced awareness of possible infection and consequently early testing and self-isolation in individuals with Delta compared to Omicron.^25^

After adjustment for potentially confounding factors and accounting for the interaction between variant and vaccination status, the risk of transmission was reduced for those with a booster dose of a COVID-19 vaccine for both Omicron and Delta. These results support early findings of vaccine effectiveness against infection with Omicron which indicates higher effectiveness among individuals with a third dose of the vaccine.^23^ The interaction observed between vaccination status and variant indicates that vaccination status, in particular a recent booster vaccination, may not have substantially decreased the number of household clusters where the index case had Omicron, whereas a booster vaccine significantly reduced household clustering arising from Delta. This indicates that viral shedding still occurs despite a booster vaccination. These findings align with initial findings from Denmark and recent data from the Netherlands. ^26 27^

This is one of the first studies to investigate Omicron transmission in households and outside of household settings among close contacts. Having a robust national level dataset and being able to link individual cases by address to secondary cases, as well as through named household contacts identified via contact tracing, have allowed for two complementary methods to assess household transmission in England. Routine collection of named contacts outside the household in national contact tracing of all cases, additionally allowed the evaluation of transmission in these settings. Through linkage of national immunisation datasets to both cases and contacts, we were able to robustly assess the reduction in transmission from vaccination, and evaluate the effect of different doses, on risk, both for onward transmission from the index, and for susceptibility of the contact.

These results must be interpreted with caution due to the potential impact of increased case finding among contacts of individuals testing positive for Omicron, although this was focused before our study period. Conversely, as the fourth wave of COVID-19 infections began and considerable demands for tests resulting in known shortages in supply, it is likely that not all clusters or transmission events were detected.^28^ Furthermore, it is possible that people were more cautious closer to the Christmas period which may have suppressed transmission rates observed. However, this is unlikely to have differentially affected Omicron compared to Delta cases. Secondary attack rates from routinely-collected contact tracing data are likely to be lower bounds due to limitations of data completeness and quality caused by variation in testing behaviour and engagement with contact tracing. Our assessment of impact of vaccine dosing on risk of transmission did not consider the timing of the vaccinations received, and as such, we could not distinguish the effect of multiple doses from recency of the vaccination.

### Conclusion

In summary, our study identified increased risk of transmission from Omicron compared to Delta, in part explained by an attenuated impact of vaccination on reducing transmission for Omicron compared to Delta. As such, Omicron’s worldwide success may be more attributable to immune escape and a milder symptom profile than increased infectivity. Our findings underscore the value of assessing growth advantage of new variants as they emerge to inform potential public health interventions such as the roll-out of the booster vaccinations and other non-pharmaceutical public health interventions.

## Data Availability

Data may be available upon request.

## Contributors

HA, ET, CT, GD, MK, CA, and TL designed the study. DC, PB, AL, CIJ, DS, FC, JF, AZ, CC, CR and GH provided data and analytical support. HA, ET, CT, MK, and AC conducted the statistical analyses. HA, ET, CT, CA, MK and TL prepared the manuscript. HA, ET, CT, CA, PB, CIJ, FC, JF, AZ, CC, CR, GH, DC, SN, ST, GD, AC, MK, and TL contributed to the writing of the manuscript.

## Acknowledgements

Data for this study are patient-level information collected as part of the care and support of COVID-19 patients and their contacts, collated, maintained and quality assured by UK Health Security Agency. DS is supported by a doctoral training grant from the Biotechnology and Biological Sciences Research Council [grant number BB/M009513/1].

## SUPPLEMENTARY FIGURES

**Supplementary Table 1A:** Adjusted risk ratios of vaccination on transmission to household contacts of cases with Omicron and Delta. Reference category is vaccinated with 2 doses + 14 days.

| Household |  |  |  |  |
| --- | --- | --- | --- | --- |
| Vaccination status | Adjusted risk ratio of vaccination status on Delta cases | p | Adjusted risk ratio of vaccination status on Omicron cases | p |
| Exposer |  |  |  |  |
| Unvaccinated | 1.19 (1.12 - 1.27) | <0.0001 | 1.00 (0.92 - 1.09) | 0.9323 |
| 1 dose + 21 days | 0.96 (0.87 - 1.06) | 0.4550 | 0.94 (0.83 - 1.08) | 0.4062 |
| 3 doses + 14 days | 0.62 (0.54 - 0.72) | <0.0001 | 0.78 (0.69 - 0.88) | <0.0001 |
| Contact |  |  |  |  |
| Unvaccinated | 1.16 (1.08 - 1.24) | <0.0001 | 0.87 (0.79 - 0.95) | 0.0015 |
| 1 dose + 21 days | 0.95 (0.86 - 1.04) | 0.2627 | 0.80 (0.69 - 0.93) | 0.0029 |
| 3 doses + 14 days | 0.68 (0.62 - 0.74) | <0.0001 | 0.88 (0.79 - 0.97) | 0.0129 |
| Adjusted as in main model for exposure date, characteristics of index cases and contacts (age group, sex), whether the contact completed contact tracing and the symptom status, region of residence, IMD and ethnicity of the index case. |  |  |  |  |

**Supplementary Table 1B:** Adjusted risk ratios of vaccination on transmission to non-household contacts of cases with Omicron and Delta. Reference category is vaccinated with 2 doses + 14 days.

| Non-household |  |  |  |  |
| --- | --- | --- | --- | --- |
| Vaccination status | Adjusted risk ratio of vaccination status on Delta cases | p | Adjusted risk ratio of vaccination status on Omicron cases | p |
| Exposer |  |  |  |  |
| Unvaccinated | 1.32 (1.06 - 1.65) | 0.0125 | 1.18 (0.97 - 1.44) | 0.1044 |
| 1 dose + 21 days | 1.28 (0.9 - 1.81) | 0.1676 | 0.9 (0.64 - 1.26) | 0.5434 |
| 3 doses + 14 days | 0.84 (0.59 - 1.19) | 0.3320 | 0.95 (0.77 - 1.16) | 0.6042 |
| Contact |  |  |  |  |
| Unvaccinated | 0.86 (0.61 - 1.21) | 0.3960 | 1.05 (0.8 - 1.36) | 0.7299 |
| 1 dose + 21 days | 1.02 (0.63 - 1.66) | 0.9218 | 0.76 (0.47 - 1.25) | 0.2829 |
| 3 doses + 14 days | 0.51 (0.39 - 0.66) | <0.0001 | 0.76 (0.61 - 0.94) | 0.0115 |

Adjusted as in main model for exposure date, characteristics of index cases and contacts (age group, sex), whether the contact completed contact tracing and the symptom status, region of residence, IMD and ethnicity of the index case.

**Supplementary Table 2.**
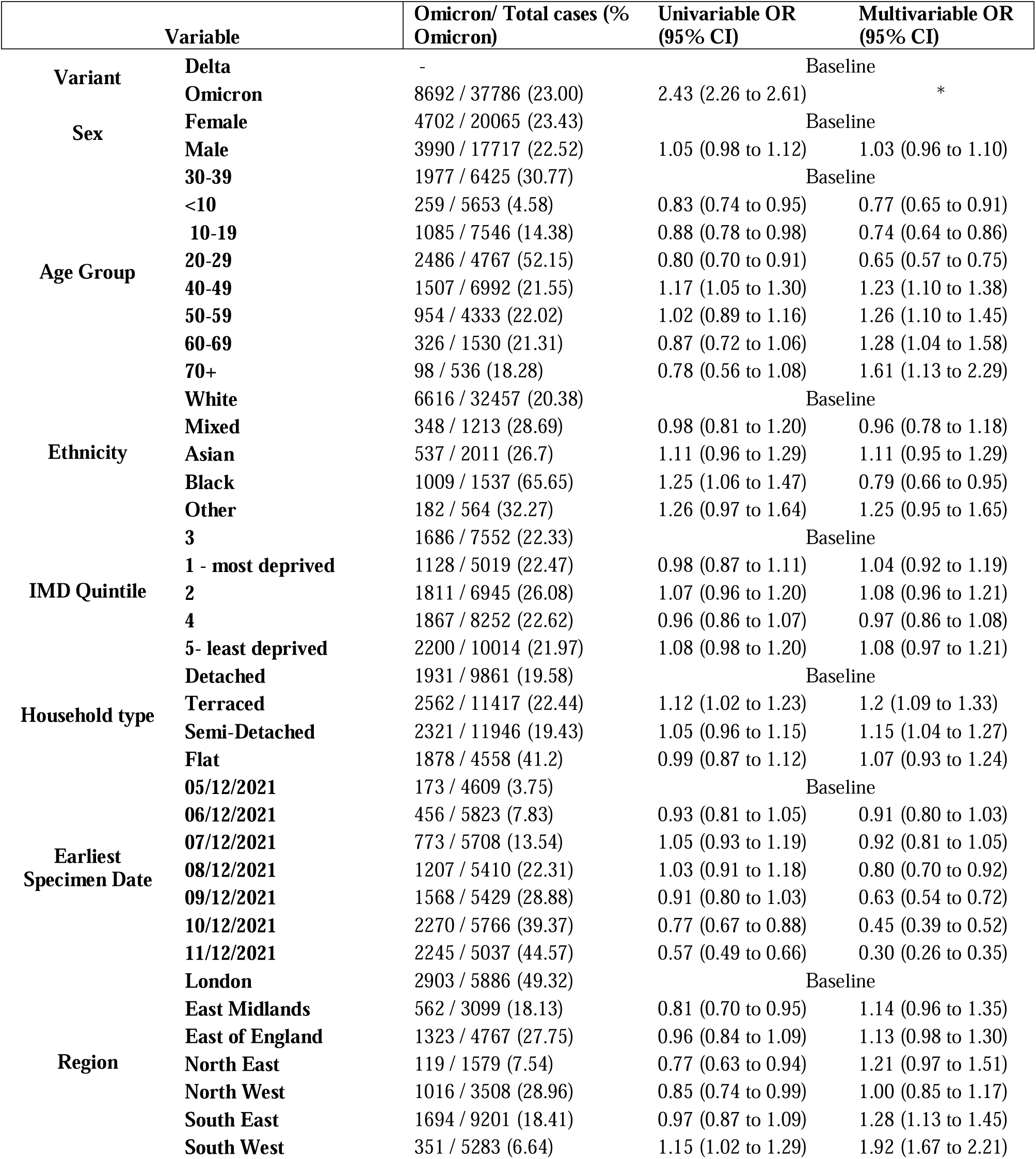
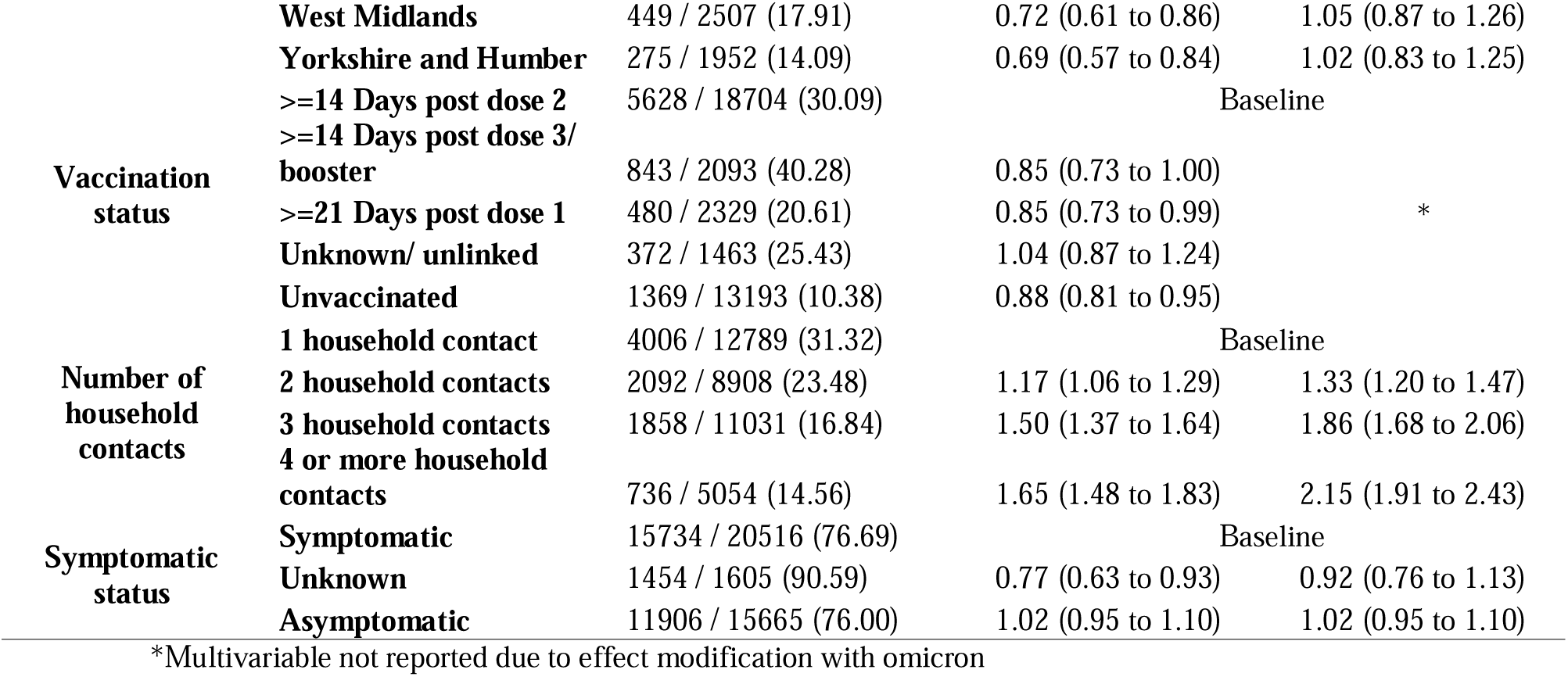

**Table S3:**
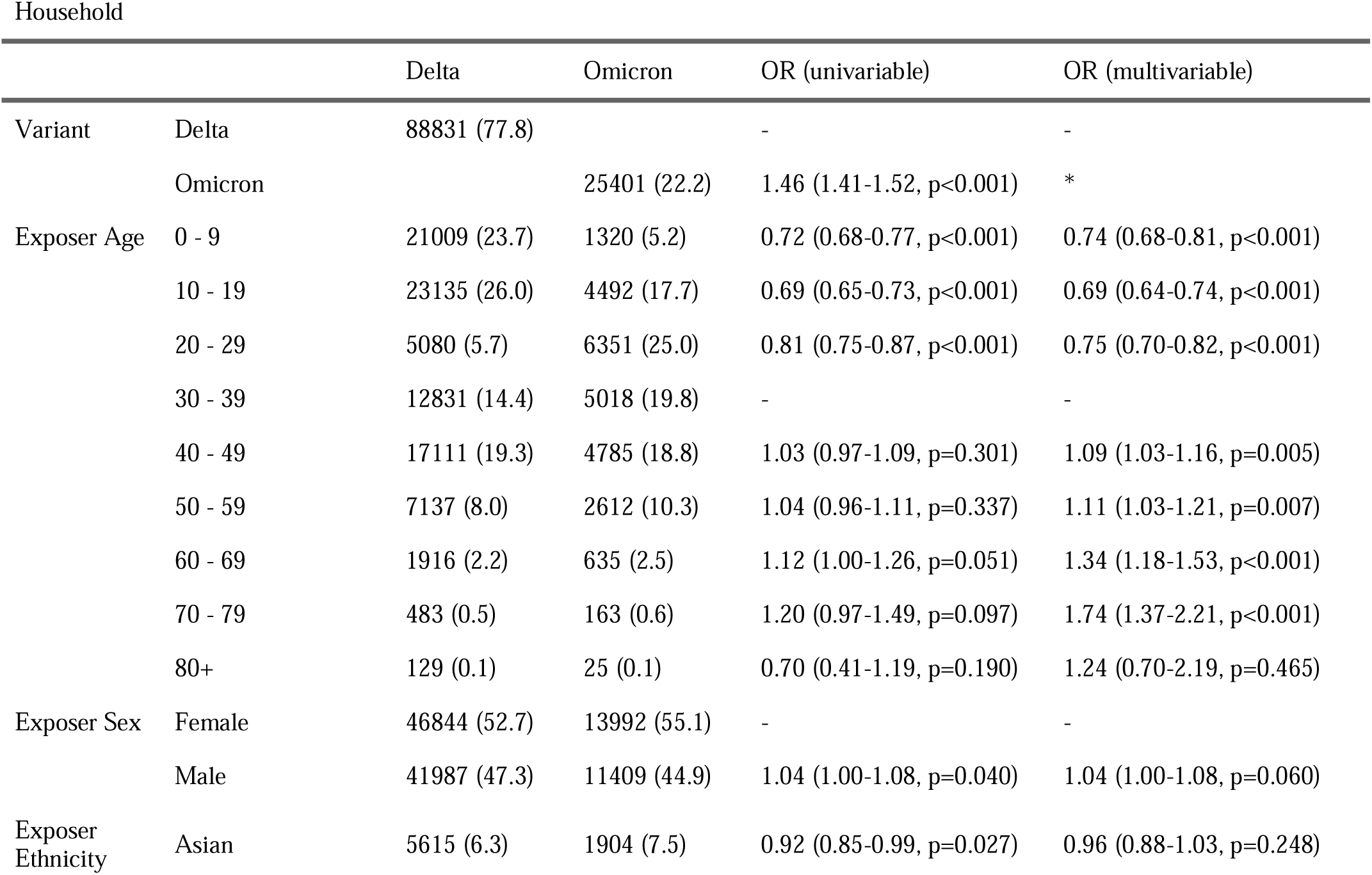
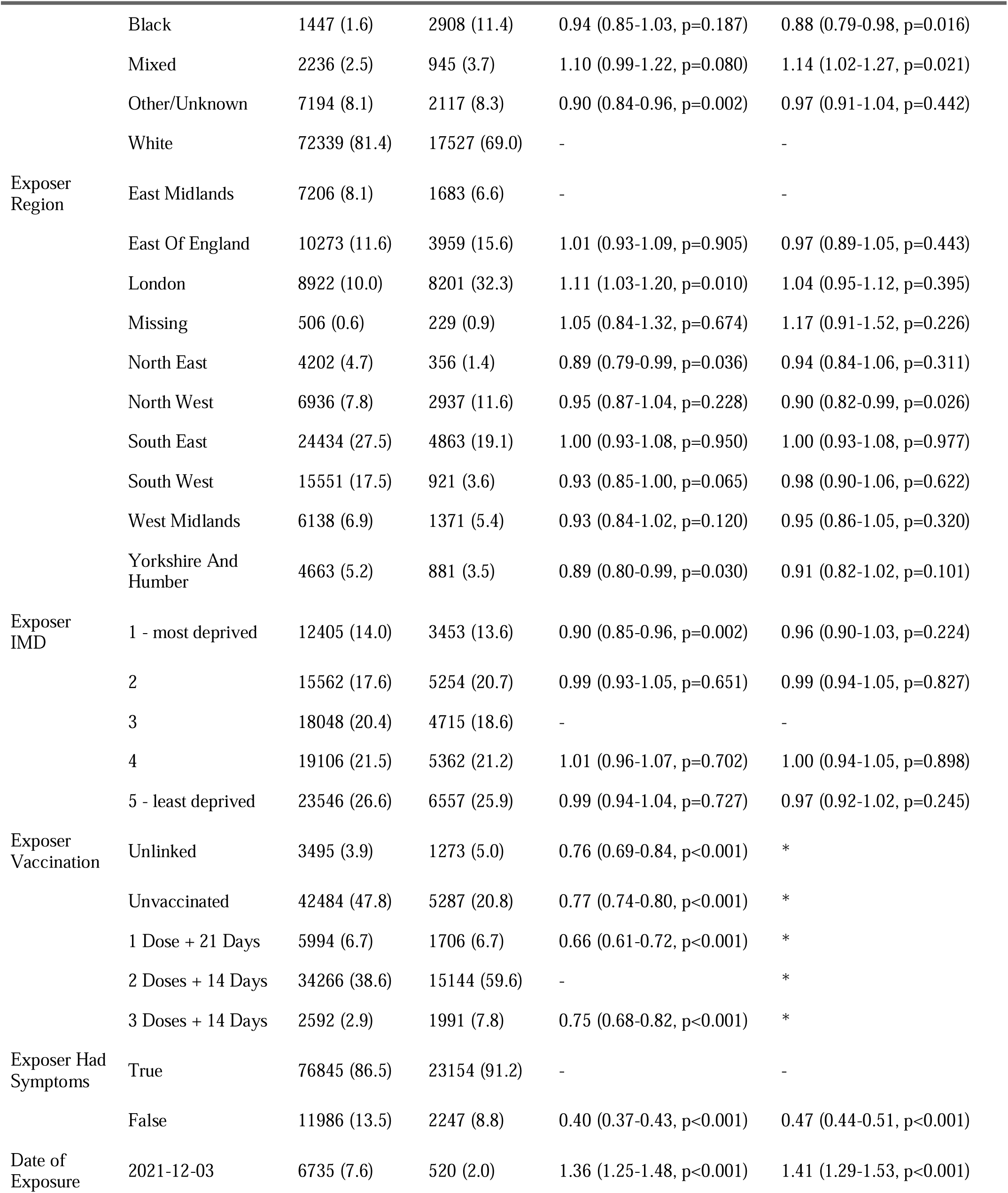
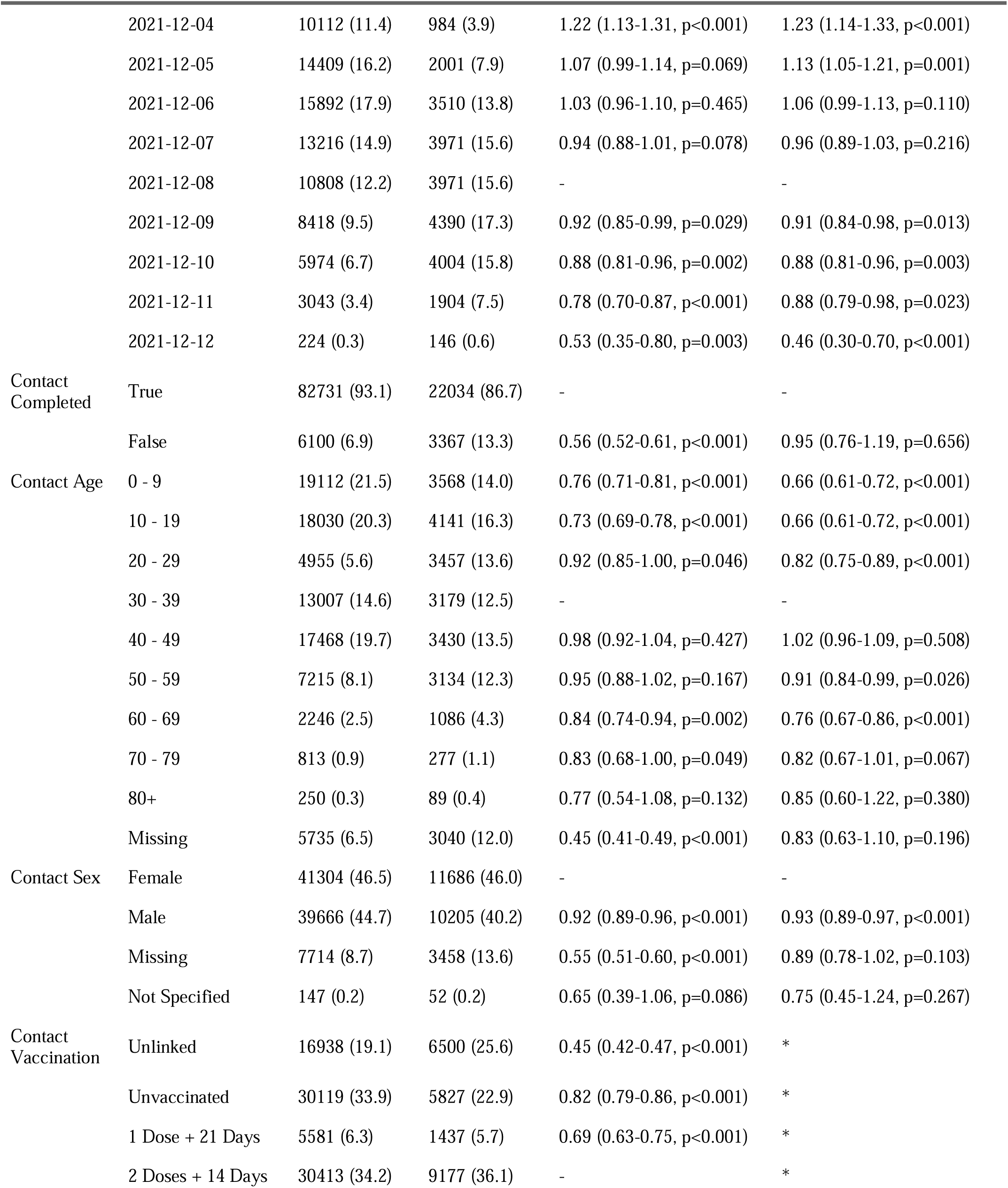
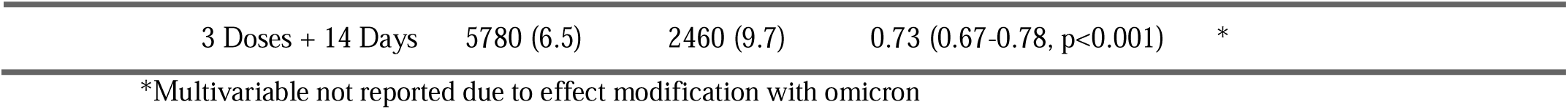
Descriptive analysis of household contacts and their exposing cases, and results of household model

**Table S4:**
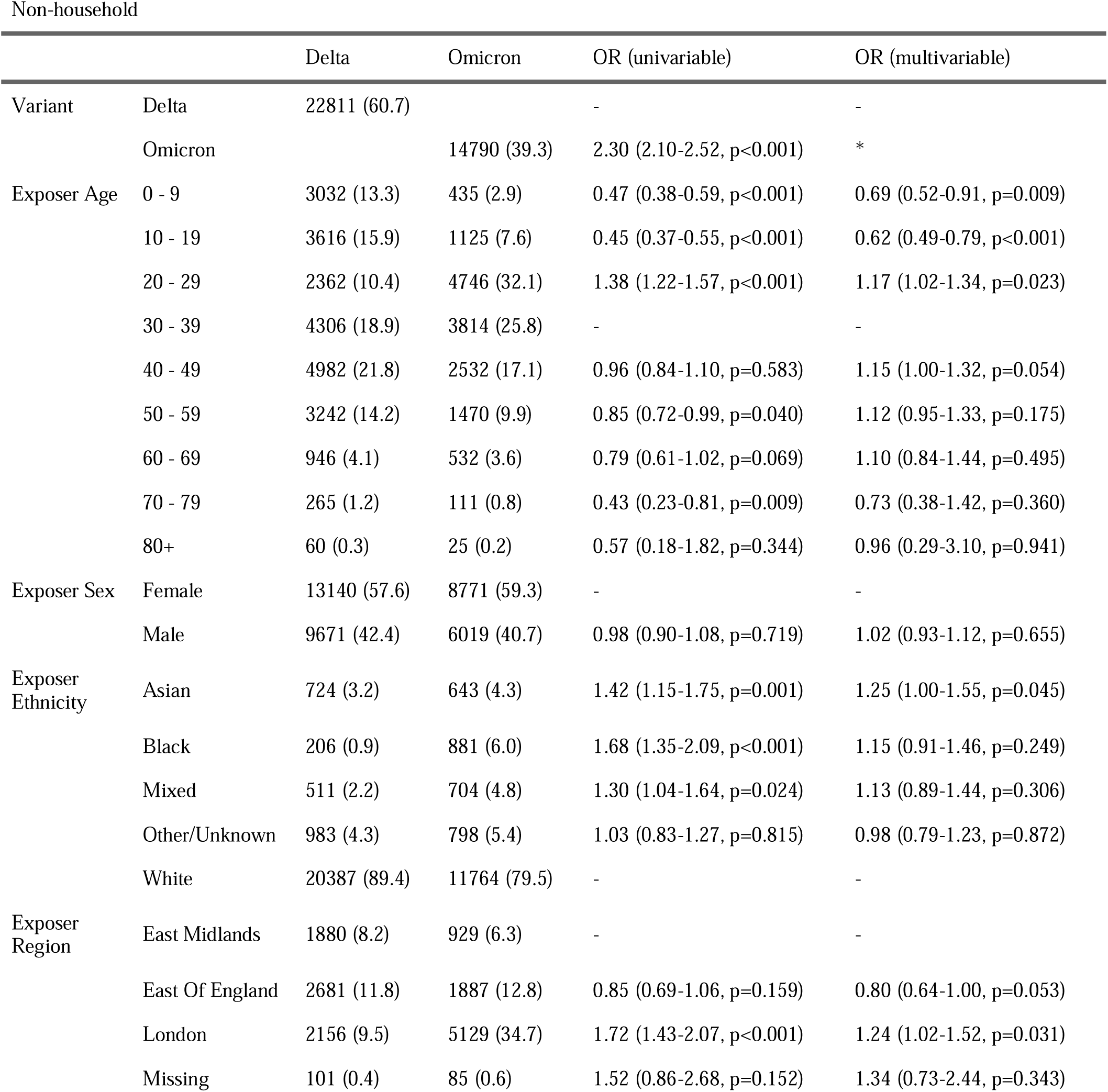
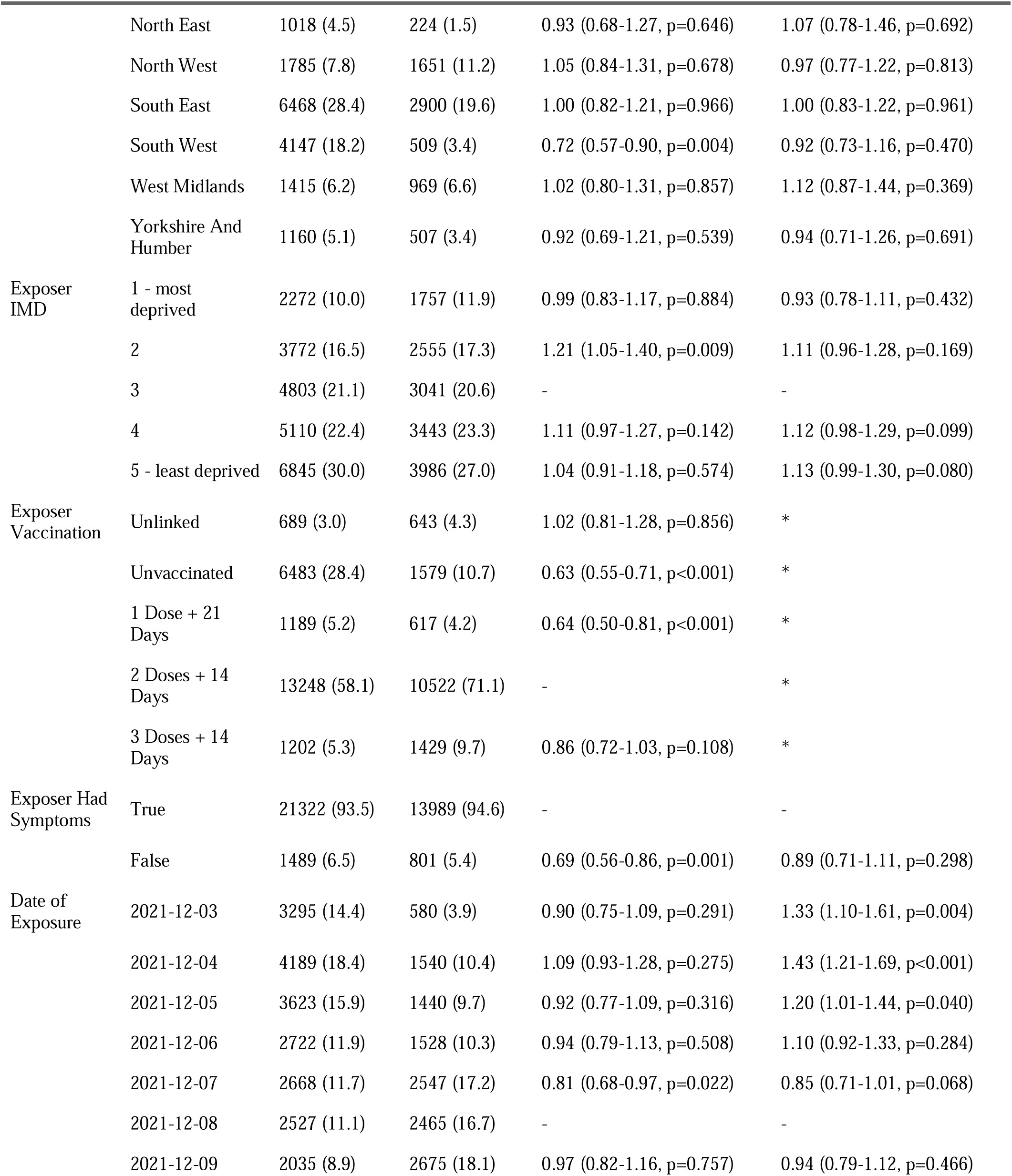
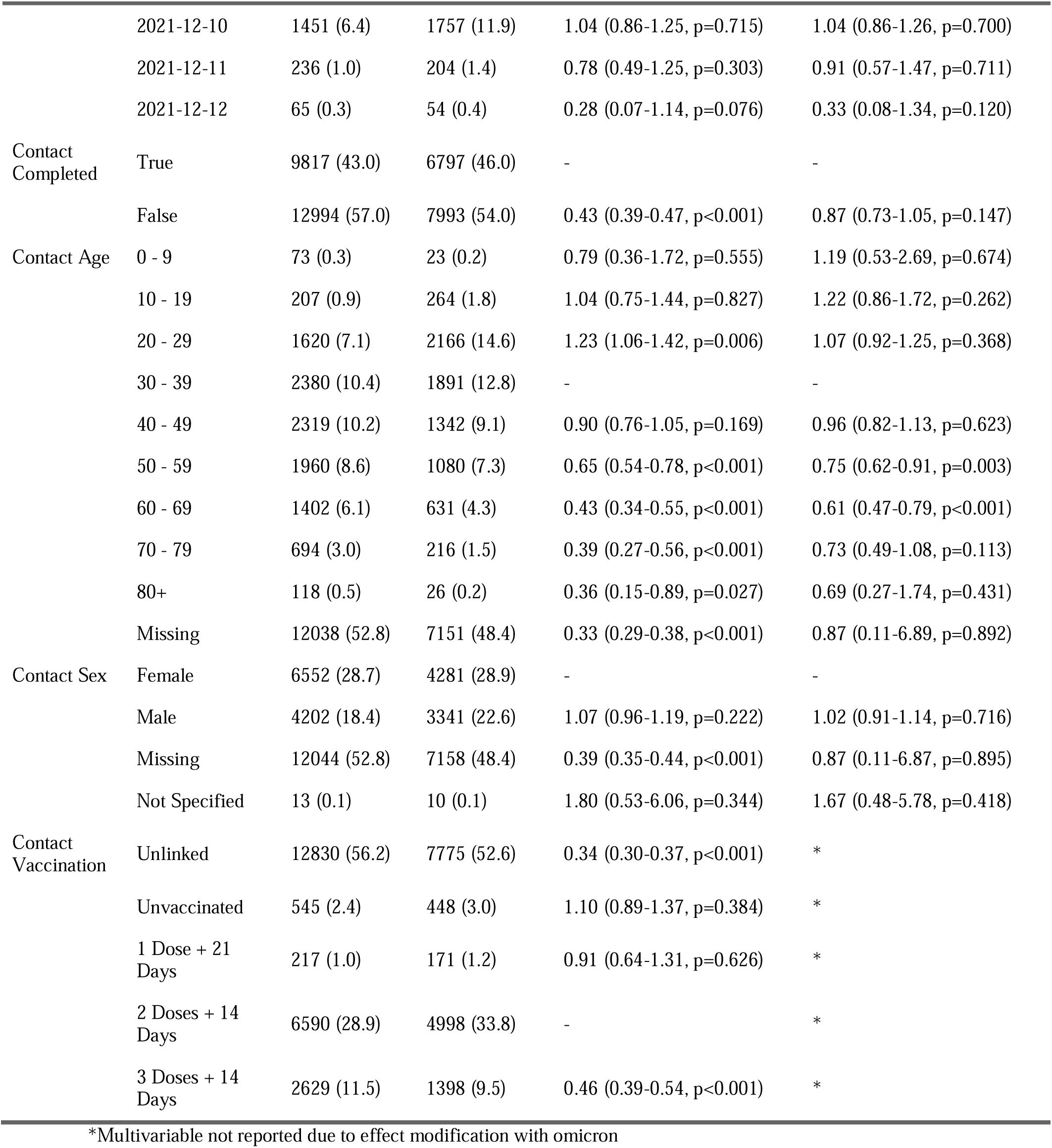
Descriptive analysis of non-household contacts and their cases, and results of nonhousehold model

